# Evaluating cotinine cutoffs and sociodemographic factors associated with exclusive heavy e-cigarette vaping behavior

**DOI:** 10.1101/2025.02.16.25322376

**Authors:** Adith S. Arun, Rishi Shah, Yuan Lu, Stephen R. Baldassarri, Harlan M. Krumholz

## Abstract

**Introduction:** Electronic cigarette (e-cig) use in previously tobacco-naïve individuals may increase the risk of nicotine addiction and adverse health outcomes, including lung injury and cardiovascular disease. This study identifies a subgroup of heavy exclusive e-cig vapers, characterizes their demographics, and establishes an optimal serum cotinine threshold to differentiate them from tobacco-naïve individuals.

**Methods:** We analyzed data from the National Health and Nutrition Examination Survey (2013– 2020) to characterize serum cotinine levels by e-cig use frequency. The optimal cotinine cutoff for identifying heavy exclusive vapers was 2.2 ng/mL (sensitivity 98%, specificity 96%).

We used receiver operating characteristic curve analysis to identify the cotinine threshold for discriminating heavy exclusive e-cig vapers from tobacco-naïve individuals and logistic regression to explore sociodemographic factors.

**Results:** Exclusive heavy e-cig vapers—defined as those who vaped on four or more of the past five days—had mean cotinine levels (220 ± 38 ng/mL) comparable to cigarette smokers (212 ± 6 ng/mL). These vapers were more likely to be 12-18 (AOR 7.96; 95% CI: [3.38 – 18.7]) or 19-25 [AOR 7.36 [2.84 - 19]) years old and less likely to be female (AOR 0.3 [0.16-0.57]).

**Conclusion:** Heavy exclusive e-cig vapers had serum cotinine levels similar to cigarette smokers and were predominantly young males. Public health interventions should target this high-risk subgroup to reduce potential long-term health consequences.

## Introduction

E-cigarettes (“e-cigs”) are used by more than 10% of high school students and 4.5% of U.S. adults. While e-cigs may promote smoking cessation in cigarette smokers, they pose risks for addiction and adverse health outcomes in previously tobacco-naïve individuals. Among all e-cig users, 16% are never smokers—representing a vulnerable group with no harm reduction benefit from vaping. This study focuses on identifying and characterizing heavy exclusive e-cig vapers, a subgroup at particularly high risk.

Health outcomes data for exclusive e-cig vapers are limited, partly because most e-cig users have concurrent cigarette smoking histories, which complicates analyses of exclusive vaping. We aim to establish a serum cotinine threshold to distinguish heavy exclusive e-cig vapers from tobacco-naïve individuals and identify sociodemographic factors associated with heavy vaping. This information may help target public health interventions and improve long-term outcome monitoring in clinical cohorts.

## Methods

We used National Health and Nutrition Examination Survey (NHANES) data from the 2013-2020 cycles^10^. Serum cotinine was used to quantify nicotine exposure. Tobacco-naïve individuals -- those self-reporting no use of any nicotine product and no second-hand smoke (SHS) exposure -- and exclusive e-cig vapers without SHS exposure were included in our analysis. We accounted for the complex survey design of NHANES to ensure nationally representative estimates. We estimated an optimal serum cotinine threshold for discriminating exclusive e-cig vapers from tobacco-naïve individuals with a receiver operating characteristic curve analysis. We used a multivariable logistic regression to determine factors associated with e-cig vaping. In self-reported tobacco-naïve individuals, concordance was defined as individuals with serum cotinine below the optimal threshold. In self-reported exclusive e-cig vapers, concordance was defined as individuals with serum cotinine above threshold. Yale University Institutional Review Board waived this study from review because it used publicly available deidentified population-level data. Data analysis was performed with R version 4.1.2. We adhered to STROBE reporting guidelines.

## Results

Of the 13,218 individuals in our sample, 179 (1.3%) were exclusive e-cig vapers. Serum cotinine levels increased, and variability decreased as the self-report frequency of e-cig use increased (Fig. 1A). Serum cotinine levels for heavy vapers (220 ± 38 ng/mL), defined as those who vaped four or more of the past five days (n=104), were similar to that of cigarette smokers (212 ± 6 ng/mL) (Fig. 1A). Serum cotinine levels distinguished tobacco-naïve individuals from light (AUC: 0.88 [0.83-0.92]) and heavy (AUC: 0.99 [0.98-0.99]) exclusive e-cig vapers (Fig. 1B). The optimal cutoff of 2.2 ng/mL for identifying heavy exclusive e-cig vapers from tobacco-naïve individuals had a sensitivity of 98% and a specificity of 96%.

**Figure 1.**
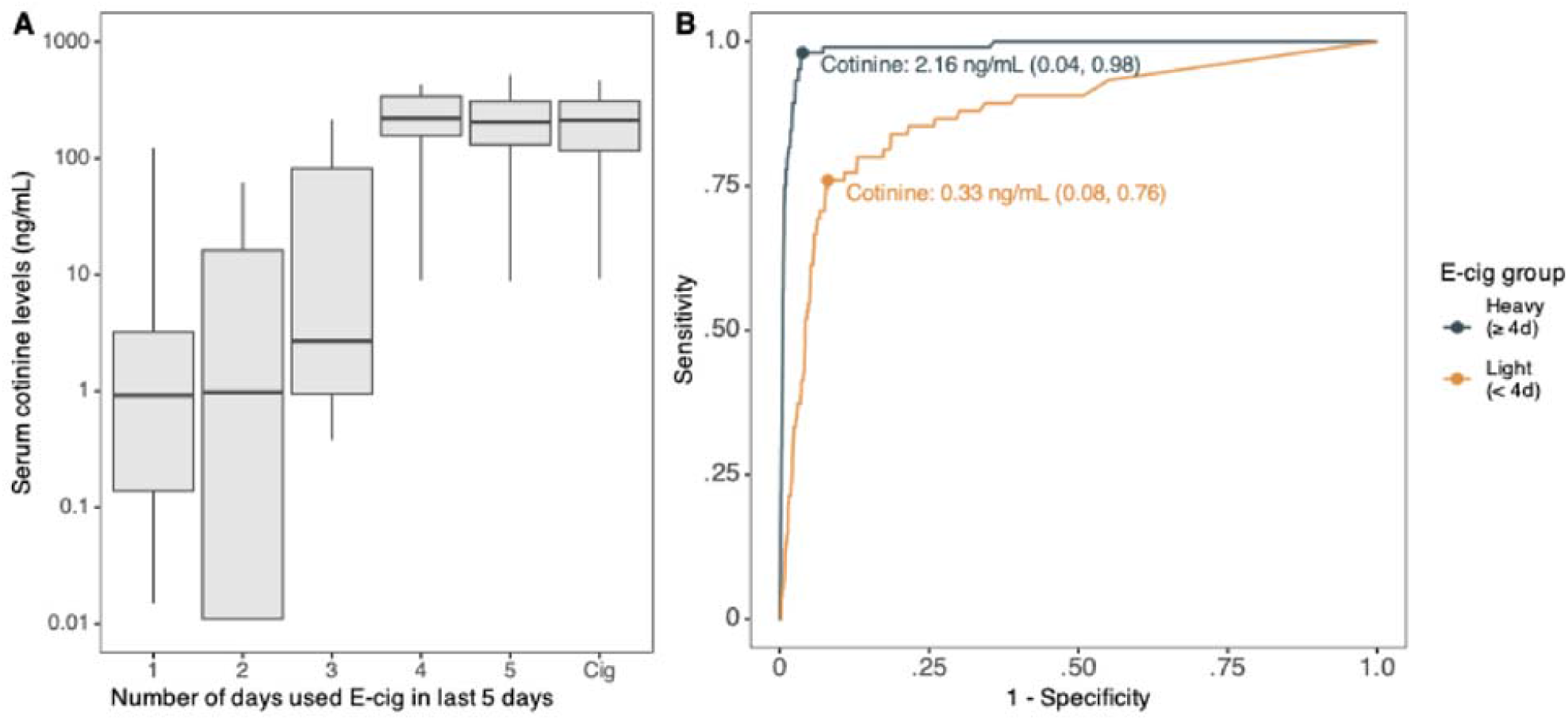
Serum cotinine levels by e-cig vaping frequency and receiver operating characteristic curve for cotinine as a marker of e-cig use. **A**) Distribution of serum cotinine levels by e-cig use frequency in the last 5 days for exclusive e-cig vapers and cigarette smokers. **B**) Receiver operating characteristic curve for distinguishing heavy (navy) and light (orange) exclusive e-cig vapers from tobacco-naïve individuals using serum cotinine levels. Optimal cotinine threshold for identifying heavy exclusive e-cig vapers from tobacco-naïve individuals was 2.2 ng/mL with a sensitivity of 0.98 and specificity of 0.96. Optimal threshold for identifying light exclusive e-cig vapers from tobacco-naïve individuals was 0.33 ng/mL with a sensitivity of 0.76 and specificity of 0.92.

Nearly all tobacco-naïve individuals and exclusive heavy e-cig vapers had self-report statuses concordant with cotinine levels (Table 1). Higher odds of exclusive heavy e-cig vaping were observed for 12-18 (7.96) and 19-25 year old (7.36) individuals (Table 1). Lower odds of e-cig vaping were observed among females (0.31), and non-Hispanic Black individuals (0.2) (Table 1).

**Table 1.** Concordance between self-reported e-cig use and serum cotinine levels, adjusted odds ratios (AOR), and sociodemographic characteristics for the US population ≥ 12 years.

| Characteristic | Tobacco-Naive Individuals<br>(*n* = 13,039)<sup>a</sup> |  | Exclusive Heavy E-Cigarette<br>Vapers (*n* = 104)<sup>a</sup> |  | AOR (95% CI)<sup>d</sup> |
| --- | --- | --- | --- | --- | --- |
|  | No. of Persons<br>(%)<sup>b</sup> | Concordance<br>With Cotinine<br>Level, %<sup>c</sup> | No. of Persons<br>(%)<sup>b</sup> | Concordance<br>With Cotinine<br>Level<sup>c</sup>, % |  |
| **Age** |  |  |  |  |  |
| ≥ 26 years | 11,389 (88%) | 98.6% | 71 (53%) | 96.8% | 1 [Ref.] |
| 19-25 years | 1,321 (11%) | 96% | 25 (42%) | 92.1% | 7.36 (2.84-19) |
| 12-18 years | 329 (1.6%) | 100% | 8 (5.6%) | 91.8% | 7.96 (3.38-18.7) |
| **Sex** |  |  |  |  |  |
| Male | 5,863 (45%) | 98.5% | 66 (72%) | 95.7% | 1 [Ref.] |
| Female | 7,176 (55%) | 97.4% | 38 (28%) | 96.6% | 0.31 (0.16-0.57) |
| **Race and Ethnicity** |  |  |  |  |  |
| NH White | 4,699 (64%) | 97.2% | 66 (86%) | 97% | 1 [Ref.] |
| NH Black | 2,525 (9.6%) | 91.3% | 10 (2.3%) | 100% | 0.2 (0.09-0.48) |
| Mexican American | 2,179 (10%) | 97.2% | 3 (1.4%) | 100% | 0.1 (0.03-0.36) |
| Other Hispanic | 1,538 (7.1%) | 97.1% | 11 (5.6%) | 100% | 0.66 (0.28-1.54) |
| Other Race | 2,098 (9.0%) | 96.9% | 14 (4.9%) | 100% | 0.41 (0.18-0.96) |
| **Income<sup>e</sup>** |  |  |  |  |  |
| High Income | 9,579 (89%) | 96.8% | 88 (94%) | 97.7% | 1 [Ref.] |
| Low Income | 2,043 (11%) | 94.3% | 10 (5.9%) | 100% | 0.58 (0.21-1.61) |

Abbreviations: NH, non-Hispanic. CI, confidence interval.
<sup>a</sup> Self-reported exclusive heavy e-cigarette vapers were defined as individuals who reported having used e-cigarettes at least four of the past 5 days. We excluded individuals who concurrently reported using any forms of smoked or smokeless tobacco or nicotine replacement products such as cigarettes, pipes, cigars, snuff, snus, dissolvables, chewing tobacco, nicotine patches, gum, lozenges, inhalers, or nasal sprays within the previous 5 days. Self-reported tobacco-naïve individuals were persons who reported not having used any of the aforementioned tobacco products or nicotine replacement products (including e-cigarettes) within the previous 5 days.
<sup>b</sup> Respondent counts are unweighted and percentage estimates are weighted; certain proportions may not sum to 100% or their total value due to incomplete/missing survey responses.
<sup>c</sup> A serum cotinine threshold of $\geq 2.16$ ng/mL was used to identify heavy exclusive e-cigarette users.
<sup>d</sup> An AOR $> 1$ indicates a higher likelihood of e-cigarette use.
<sup>e</sup> Low income was defined as a respondent's ratio of family income to the federal poverty threshold for the year of interview being $\leq 1$

## Discussion

We identified a subgroup of heavy exclusive e-cig vapers with serum cotinine levels nearly identical to those of cigarette smokers. Our findings suggest that serum cotinine is a reliable marker for distinguishing heavy exclusive e-cig vapers from tobacco-naïve individuals, with an optimal threshold of 2.2 ng/mL. This threshold offers a valuable tool for future research and clinical screening to monitor nicotine exposure and associated risks in young, previously nicotine-naïve populations.

The demographic profile of heavy exclusive vapers highlights that young males are at the highest risk for sustained nicotine exposure through e-cig use. In contrast, non-Hispanic Black individuals were less likely to be exclusive e-cig vapers, which may reflect differences in e-cig use patterns or socioeconomic factors influencing access and preferences for nicotine products. Notably, non-Hispanic Black individuals were less likely to be exclusive e-cig vapers. Previous work considering both exclusive and non-exclusive e-cig vapers estimated 30% of e-cig vapers were non-Hispanic Black, in contrast to our national estimate of 10% reflecting the possibility that non-Hispanic Black e-cig vapers may be preferentially using e-cig vapes as nicotine replacement or concurrently with other modes of nicotine delivery^9^. The small sample size and potential biases in self-reported vaping behavior warrant further validation studies in larger cohorts.

Efforts to mitigate the risks associated with heavy exclusive e-cig use should focus on early intervention, particularly in adolescent and young adult populations. The long-term health outcomes of heavy exclusive e-cig vaping are largely unknown but linked to cardiovascular disease risk, e-cig associated lung injury, and addiction^5-7^. Serum cotinine monitoring may complement self-reported data in clinical practice to ensure accurate identification of high-risk individuals. Future studies should explore how this biomarker threshold can be integrated into public health surveillance systems and linked to long-term health outcomes.

## Data Availability

All data produced in the present work are contained in the manuscript and further requests are available upon request to the authors.

